# COVID-19 vaccine hesitancy in care home staff: a survey of Liverpool care homes

**DOI:** 10.1101/2021.03.07.21252972

**Authors:** John S P Tulloch, Karen Lawrenson, Adam L Gordon, Sam Ghebrehewet, Matthew Ashton, Steve Peddie, Paula Parvulescu

**Affiliations:** Institute of Infection, Veterinary, and Ecological Sciences, University of Liverpool, Liverpool, UK; Public Health Department, Liverpool City Council, Liverpool, UK; School of Medicine, University of Nottingham, Nottingham, UK; Public Health England North West, Cheshire & Merseyside Health Protection Team, Liverpool, UK; Adult Social Care Department, Liverpool City Council, Liverpool, UK

**Keywords:** COVID-19, care homes, vaccine hesitancy

## Abstract

**Background:** COVID-19 has caused high morbidity and mortality in UK care homes. Vaccinating staff members and residents will protect care homes from severe clinical cases. Uptake of COVID-19 vaccine first doses in care homes has been higher among residents compared to staff members.

**Methods:** We aimed to identify causes of lower COVID-19 vaccine uptake amongst care home staff members within the Liverpool City Council region. An anonymised online survey was distributed to all care home managers between the 21^st^ and the 29^th^ of January 2021. Descriptive analysis was performed on responses.

**Results:** 46/87 (53%) of Liverpool care homes responded. The mean staff vaccination rate per home was 51.4% (95% CI 43.9-58.8%). The most common reasons for staff not receiving the vaccine were: concerns about lack of vaccine research (37.0%), staff being off-site during vaccination sessions (36.5%), pregnancy and fertility concerns (5.6%), and concerns about allergic reactions (3.2%). Care home managers reported the necessity to combat vaccine hesitancy through meetings and conversations with health professionals, and provision of supporting evidence to dispel vaccine misinformation.

**Conclusions:** Vaccine hesitancy was the main cause for reduced vaccine uptake among care home staff members. These concerns could be addressed by targeted evidence-based training, and a public health communication campaign to build vaccine confidence and increase acceptance of COVID-19 vaccines. The speed of vaccination roll-out has also led to unexpected logistical issues that lowered vaccine uptake rates. Addressing both these challenges could increase uptake by more than 40%.

**Key Points:**

- COVID-19 vaccine uptake rates are lower in staff than residents
- Three main causes of reduced uptake have been identified: vaccine hesitancy, logistical issues, and medical concerns.
- The main reasons for vaccine hesitancy were concerns about limited research into vaccine safety, and concerns about long-term impact on pregnancy and fertility.
- Addressing care home staff vaccination concerns should be given priority in these settings.

## Introduction

Since the start of the COVID-19 pandemic, 23.8% of all care home deaths have been due to COVID-19 [1]. The majority of these occurred in homes which had experienced a COVID-19 outbreak [2]. The Liverpool City Council (LCC) region had significantly more COVID-19 related deaths in its care home population (31.5%, n=224) compared to the national average; a risk ratio of 1.33 (95% 1.19-1.48, p<0.001) [1]. At least 62% of Liverpool care homes have experienced COVID-19 outbreaks [3]. LCC serves a population of almost half a million people, and is one of the most deprived local authorities in England, with lower than average life expectancy; 14.6% of its population is over 65 [4,5].

Care home residents have high levels of frailty and multi-morbidity [6]. They are affected by immunosenescence [7], which makes them very susceptible to SARS-CoV-2 infection. There are three main portals of entry for SARS-CoV-2 into a care home: newly admitted or readmitted residents; staff; and visitors. Strategies to limit infections and outbreaks have included: improved Infection Prevention and Control (IPC); testing staff, visitors and residents; isolation and zoning; limiting non-essential professional visits; and restricting indoor visiting [8]. Despite these measures, COVID-19 outbreaks have continued [1]. The COVID-19 vaccine programme brings hope to the care home staff, residents and the wider community. Successful vaccination of care home staff and residents should result in less severe outbreaks with reduced morbidity and mortality. In order to improve population protection, it is critical that vaccine uptake amongst care home staff and residents is optimised.

International surveys have shown that 28% of the general population are COVID-19 vaccine hesitant, with the highest rates in the 25-34 age group and in females [9]. Hesitancy reasons include concerns about safety, lack of effectiveness, and the belief that vaccination is unnecessary [10]. Twenty-nine percent of health care works are hesitant, with higher levels in young adults and females, and 41% of those hesitant have safety concerns about the vaccine[11]. An American study of 11,460 care homes found only 37.5% of staff members had received a COVID-19 vaccine, compared to 77.8% of their residents [12]. Only one study has investigated COVID-19 hesitancy levels in care home staff (in Indiana, USA) [13]. In this study, 36% were reluctant, with the main barrier being concerns about side effects. Hesitancy levels were higher in female and younger members of staff.

On the 23^rd^ of December 2020 the first doses of COVID-19 vaccines were offered to care home residents and staff in the 87 care homes within LCC. By the 29^th^ of January 2021, 70.3% of care home residents, and 39.8% of staff had received their first vaccination. A rapid evaluation of the vaccination roll-out was performed to assess whether low levels of vaccine uptake in Liverpool care home staff were due to high levels of vaccine hesitancy, or other unidentified factors.

## Methods

An anonymous online survey was distributed, between the 21^st^ and 29^th^ of January 2021, to care home staff managers whose care homes (n=87) lie within the LCC region. Information was collected about the number of permanent staff employed at the home and the number of staff that had not been vaccinated. Reasons for staff remaining unvaccinated were identified and the number of staff associated with each reason were quantified. Respondents [care home managers] were asked to describe what they had done to encourage vaccine hesitant staff to get vaccinated and what further assistance they required. All data collated from the survey were analysed descriptively.

## Results

Fifty-three percent (52.8%, n=46) of care home managers in Liverpool responded with results available for analysis. In total, these homes employed 2128 individuals, with a median staff size of 38 (range:1-166). The overall COVID-19 first vaccination rate reported by staff was 52.6% (n=1119), with a mean vaccination rate per care home of 51.4% (95% CI 43.9-58.8%) (Fig 1).

**Fig 1.**
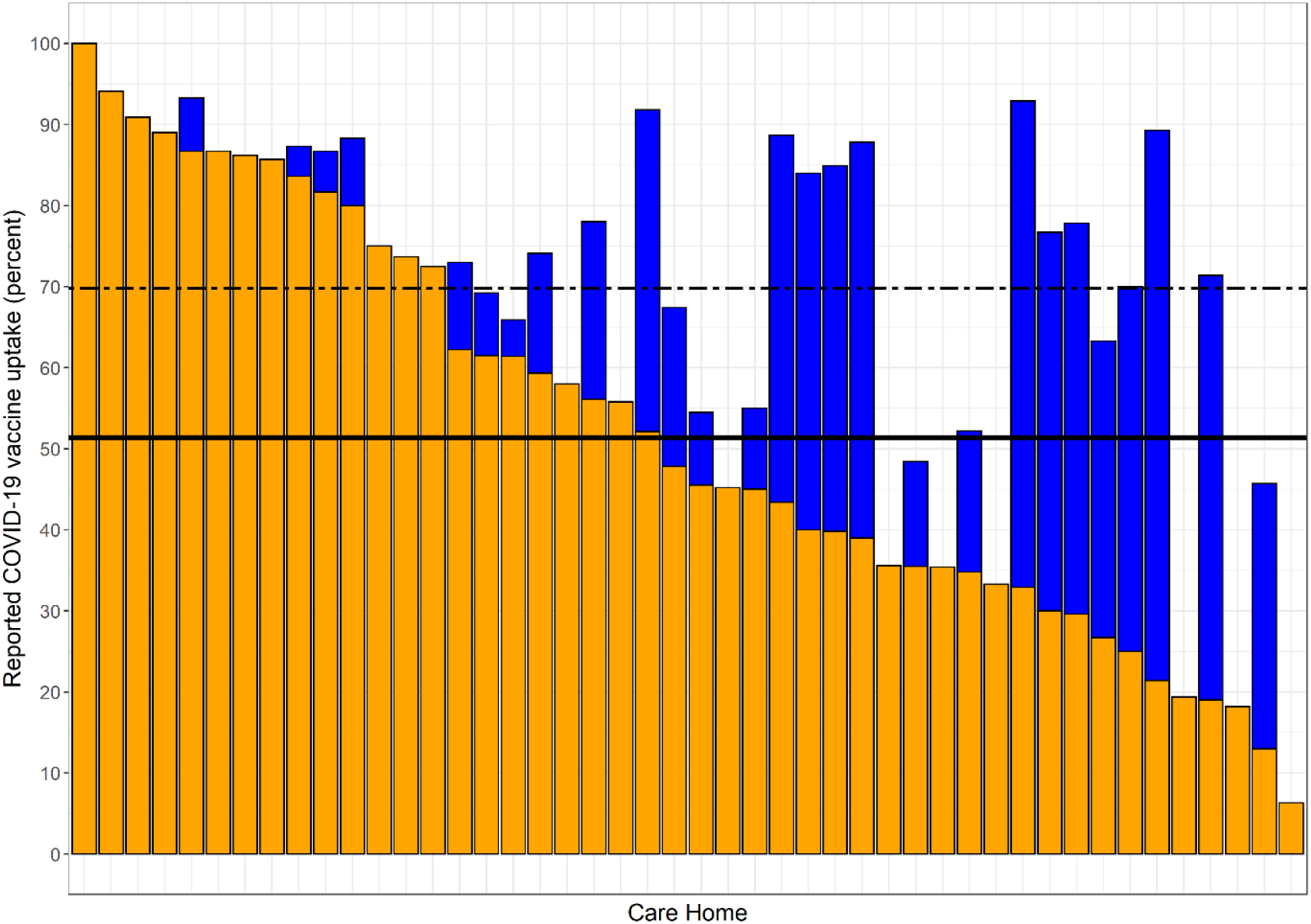
Vaccination uptake rate in Liverpool care home staff. Orange columns represent the self-reported vaccine uptake rates in each home. Blue columns represent potential vaccine uptake rate if only logistically issues are resolved. The solid black line represents the mean vaccine uptake rate. The dashed black line represents the predicted mean vaccine uptake rate if logistical issues are resolved.

Twenty four percent (24.2%) of care home staff were not vaccinated due to vaccine hesitancy, 18.4% due to logistical issues, and 4.2% due to medical concerns (Table 1). The belief that not enough research had been performed into vaccine safety was present in almost all homes (82.6%). Logistical issues impacted over half of care homes. If logistical issues were resolved, the mean vaccination rate could have increased to 69.8% (95% CI 63.2-76.3%) (Fig 1). Health concerns were widespread and were prevalent reasons for not receiving the vaccine. The following fears were reported: the vaccine affecting fertility; vaccine immunity being short-lived; one could still become sick, or die, despite being vaccinated; and concerns that vaccinations would not stop transmission.

**Table 1.** Reasons for care home staff members being unvaccinated against COVID-19

| **Reported reasons for staff remaining unvaccinated** | **Proportion (95% CI) of unvaccinated staff (n=1009)** | **Proportion (95% CI) of care homes with this reason present (n=46)** |
| --- | --- | --- |
| **Vaccine Hesitancy** |  |  |
| Staff member believes that not enough research has been performed into vaccine safety | 37.0%, (34.0-40.0) | 82.6%, (69.6-91.6) |
| No reason provided | 9.4%, (7.7-11.3) | 34.8%, (22.1-49.3) |
| Staff member believes that the vaccine contains microchips | 1.9%, (1.2-2.9) | 10.9%, (4.1-22.5) |
| Staff member is afraid of needles | 1.5%, (0.9-2.4) | 13.0%, (5.5-25.2) |
| Staff member believes that the vaccine could alter their DNA | 1.4%, (0.8-2.3) | 6.5%, (1.7-16.7) |
| **Logistical Issues** |  |  |
| Staff member was not on site on the day of vaccination | 36.5%, (33.5-39.5) | 52.2%, (37.8-66.3) |
| Staff member was not offered the vaccine | 2.5%, (1.6-3.6) | 8.7%, (2.8-19.7) |
| **Health Concerns** |  |  |
| Staff member was pregnant, planning a family, or concerned about long term fertility impact of vaccine | 5.6%, (4.3-7.2) | 43.5%, (29.8-58.0) |
| Staff member has been advised by health professionals not to be vaccinated due to a history of allergic reactions | 3.2%, (2.2-4.4) | 34.8%, (22.1-49.3) |
| **Health-related contraindications** |  |  |
| Staff member currently COVID-19 positive | 1.1%, (0.6-1.9) | 6.5%, (1.7-16.7) |

Reported methods to address vaccine hesitancy included: one-on-one meetings to discuss concerns (34.8% of care homes, n=16); staff meetings (15.2%, n=7); provision of educational material (15.2%, n=7); individual discussions with general practitioners or the vaccination team (10.9%, n=5); managers leading by example and encouragement (6.5%, n=3); and reviewing employment law to see whether vaccination could be enforced (2.2%, n=1).

Twenty-six percent (n=12) of care home managers did not want assistance in reducing vaccine hesitancy. The remainder would have liked: health professionals’ advice (e.g. forums, one-on-one calls, weekly meetings) (15.2%, n=7); information about the vaccine, including expected side effects (10.9%, n=5); ‘myth-busting’ material, especially about long-term fertility impact (6.5%, n=3); repeat visits by the vaccination team (2.2%, n=1); a local awareness campaign (2.2%, n=1); and making vaccination compulsory for care home staff (2.2%, n=1).

## Discussion

Our evaluation highlights that vaccine hesitancy and logistical challenges are the main reasons for reduced vaccine uptake amongst care home staff. Conspiracy theories about vaccines were not prevalent or widespread amongst this group of staff. The reported vaccine uptake rate of 51.4% at the date of this survey is concerning. This is comparable to COVID-19 vaccination in American care homes [12].

The social care workforce is predominately female (82%, compared to 47% in the economically active population), and with a higher proportion of BAME individuals (21% vs 14% in England) [14]. This is a similar demographic to the parts of the general population with high levels of COVID-19 vaccine hesitancy [15,16,17]. Concerns about the lack of adequate research into vaccine safety were widespread and were the most prevalent reason for non-vaccination. These mirror concerns of the general population [15,16,17]. Strategies to quell these specific fears need to utilise personal experience alongside expert advice, in order to be successful [19]. This could include material about vaccine development, safety profile, and the number of participants in vaccine trials [18,19]. To reduce vaccine hesitancy for all vaccines, staff knowledge and awareness around general vaccine development and licensing process requirements could be improved through training.

The national COVID-19 vaccination roll-out has been a great success in the UK, but logistical issues resulted in Liverpool’s care homes having reduced vaccine uptake. On the assumption that these issues were separate from vaccination hesitancy, then, if resolved, vaccine uptake among staff members would have increased by almost 20%.

Health-associated concerns represented the smallest contributors to reduced vaccine uptake, but were often founded on erroneous information and could be easily addressed. Pregnancy and fertility associated concerns were widespread. Both vaccines’ safety briefs have limited information on this topic [20,21]. The UK government advice is that those who are pregnant and are ‘at very high risk of catching the infection or those with clinical conditions that put them at high risk of suffering serious complications from COVID-19 should be vaccinated [22].’ Care home staff members would fit within this category and should be encouraged to get vaccinated following a risk assessment. The ‘history of allergies’ reason was present in around a third of homes. Vaccine-induced anaphylaxis is an extremely rare event, and care home staff should be reassured that this is an unlikely occurrence (1.3 cases per million doses) [23]. It is important for vaccinators to be clear with staff that “history of allergies” is not the same as “history of anaphylaxis”. Emerging data from Moderna and Pfizer suggest that their vaccines have had an anaphylaxis rate of 2.5 and 11.1 cases per million doses respectively [24,25].

Conspiracy theories were not commonplace and only mentioned in a small number of care homes. This is good news, because conspiracy theories are more likely to affect the attitudes of people with neutral feelings towards vaccination; campaigns may be better targeted towards “fence-sitters” [26]. Strategies should not rely solely on directly debunking false information, but encourage engagement with health professionals, and the use of publicly visible campaigns that build vaccine confidence and encourage participation through peer pressure.

## Limitations

The survey describes self-reported rates, and views were compiled by one senior member of the care home. It is possible that this may not reflect the views of all staff members. We do not know how representative the views are of care home staff in Liverpool, nor the wider UK care home staff population. Additionally, the reported vaccine uptake rates (52.6%), were higher than what was provided through the NHS vaccine tracker to LCC (39.8%) at the time of the survey. Either care home managers were overestimating uptake, or the tracker did not provide the most up to date information.

## Conclusions

The public health emergency and severe consequences of COVID-19 in care homes has led to the rapid administration of vaccines within the care home resident and staff populations – which is an incredible success story. The necessary speed of roll-out has resulted in missed vaccinations due to last minute appointments, and vaccine-related fears could not always be allayed. This work has shown that most vaccine hesitancy in care home staff is not due to conspiracy driven theories, but due to perceived lack of adequate research into vaccine safety. These reasons could be countered by a multifaceted public health campaign, aimed at both care home staff and the wider public, to emphasise the overwhelming vaccine acceptance in the general population.

## Data Availability

Data is available upon reasonable request to Liverpool City Council

## Acknowledgements

We would like to acknowledge all care home staff who continue to provide incredible care and support to their residents during the most difficult of times.

## Ethics Statement

These data were collected as part of routine public health service evaluation by Liverpool City Council. Fully anonymised data were provided to JT for secondary data analysis. As such, the University of Liverpool ethics department confirmed that review by the University of Liverpool research ethics committee was not needed (see http://www.hra-decisiontools.org.uk/research/docs/DefiningResearchTable_Oct2017-1.pdf).

## Data Availability Statement

Data are available upon reasonable request to Liverpool City Council.

